# Understanding the mechanisms behind rotational and localized lockdowns: How mobility and spatial disparities modulate COVID-19 transmission

**DOI:** 10.1101/2025.05.02.25326869

**Authors:** Mauricio Santos-Vega, Jaime Cascante Vega, Felipe Aramburo Jaramillo, Catalina Gonzalez-Uribe, Juan Manuel Cordovez

## Abstract

City-level curfews and other non-pharmaceutical interventions (NPIs) targeting specific communities or finer within-city aggregation, such as neighborhoods or localities, were used to slow the transmission of SARS-CoV-2 upon the announcement by the World Health Organization of the emergence of a virus with a potential pandemic threat. Community transmission was reduced with coarser NPIs, but the effectiveness of NPIs restricting finer spatial scales remains to be refined, especially in big urban centers with heterogeneous spatial distribution in their mobility and socioeconomic variables. Communities are segregated spatially in large urban centers based on socioeconomic status^1–4^. In developing countries, the lowest socioeconomic statuses must commute to far locations the most^5^and sustain their economies based on informal activities^6^. Consequently, it is unclear what spatial aggregation is appropriate to intervene while minimizing economic disruption. This work evaluates an NPI implemented in Bogotá, a megacity in Latin America in Colombia. A set of rotational lockdowns at the locality scale (an administrative aggregation of neighborhoods) was implemented in the city. First, we use mobility data to investigate how commuting changed at the scale of the restrictions. Second, using population epidemiological surveillance data of COVID-19, we quantify community transmission at the scale of the restrictions by estimating the effective reproductive number *R*_*eff*_ and investigating if changes in mobility correlate with those in transmission. Third, we use an epidemiological transmission model to simulate counterfactual scenarios in the absence of NPIs. We compared the counterfactual projections with the reported incident infections, mortality, and community transmission at the city scale. Thus, we will determine whether the finer NPIs reduced the level of community transmission in the city. Finally, we use socioeconomic data at the scale of the restrictions (localities) to investigate how these and mobility predicted transmission. We incorporate random variability across space to simulate uncontrolled sources of variability. Our results show how finer NPIs change mobility at different spatial scales. Although the intervention reduces mobility between units, the finer-scale mobility is not perturbed. We have evidence that the first sets of interventions were the most effective in reducing transmission. Finally, we found substantial spatial heterogeneity in the combined effect of socioeconomic variables and mobility in predicting transmission.

## Introduction

Following the report of the first person who tested positive for SARS-CoV-2 infection on March 6th, 2020, a country-level lockdown was implemented in Colombia starting on March 20th. This country’s lockdown prohibited commuting between municipalities (n=1122 in 2020) and restricted mobility within cities^7^. This restriction reduced SARS-CoV-2 transmission, delayed outbreaks in cities across the country, and the importation of cases to naive populations^8^. In Bogotá, the capital district, which had 8 million inhabitants in 2020, the first two months after the first imported case, fewer than 100 cases were reported daily. In May 2020, the first relaxation of this country-level NPI followed an exponential increase in cases^7^ (Figure 1A). In response to the rise in COVID-19 infections, interventions nationwide were designed and implemented at finer scales to reduce transmission and prepare the healthcare system for the expected burden. In Bogotá, these finer NPIs included limited access to food supply markets and grocery stores, restricted use of public spaces, closure of schools and universities, and mask use^9^. Essential workers involved in manufacturing and construction were allowed to attend their workplaces and maintain their pre-pandemic schedules. Despite these control measures partially restricting mobility, COVID-19 cases continued to increase^10^. By July 20th, the ICU exceeded 80% of its capacity, and to reduce the economic impact of another city-level lockdown, finer NPIs were implemented at the locality level. Localities are defined as administrative aggregations, and most people are aware of the ones they live in. This aggregation spans various spatial areas and has heterogeneous population densities, among other variables (Supplementary information SI Figure S1). Indeed, public transportation is sectorized at this spatial scale, and reductions in mobility during the first month of the pandemic assorted at this scale ^11^. Therefore, these finer NPIs were planned by the major’s office COVID-19 response team and included a varying timing of curfews at an aggregation of localities (Figure 1C). These NPIs were implemented as a rotational lockdown (across the city) scheme at this spatial unit. Localities were selected based on the previous week’s cumulative reported infections. Four sequential NPI rounds were deployed between July and September to control the spread of SARS-CoV-2 in Bogotá (see Figure 1A, C for a schematic of the interventions). In this work, we combine epidemiological surveillance, mobility, and socioeconomic data to evaluate the impact of these interventions on community transmission and the changes in the number of infections and mortality due to COVID-19.

**Figure 1.**
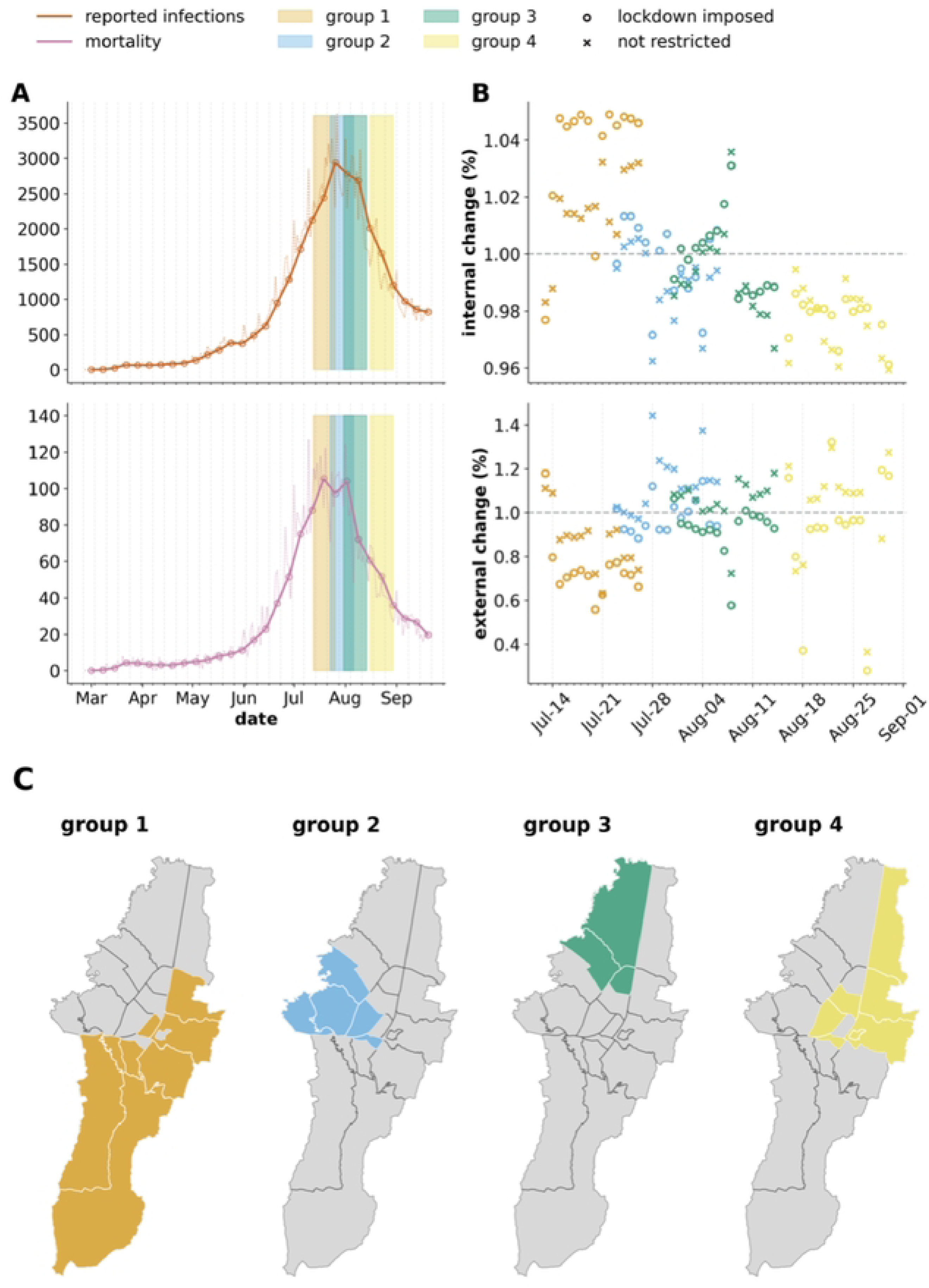
Diagram showing the timing of each lockdown in Bogotá, highlighting the epidemiological surveillance and mobility. In A), we present the daily incident reported infections (upper plot) and reported deaths (lower plot) as dashed lines, and we also present the weekly average as a continuous line. We include ribbons to schematically show the regions of the rotational lockdowns. In B), we show how both internal (upper plot) and external (lower plot) mobility changed in localities with imposed restrictions compared with mobility before restrictions at their respective spatial scales. The colors correspond to the lockdown group, and we highlight differences in the localities under a restriction (circles, o) and those without interruption (crosses, x). In C), we highlight the localities and color them under a restriction in each group, as indicated on the upper left side of each map. All the colors are consistent across the plots.

The rapid spread of SARS-CoV-2 globally unchained the worst pandemic in human history in the past century. Non-pharmaceutical interventions (NPIs) were implemented worldwide at various spatial scales to control the spread of the virus ^12,13^. Because individuals’ immune systems are entirely naive to an emerging virus without previous exposure, social distancing and lockdown measures are the first available control tools^14,15^. Therefore, when SARS-CoV-2 likely emerged in the seafood market in Wuhan, China^16,17^, lockdowns controlled its spread within communities, prevented the overflow of healthcare systems, and reduced mortality^18^. These interventions were implemented to slow transmission and wait until the development of pharmaceuticals to prevent infections and reduce transmission and the probability of disease^19^. However, quantifying the impact of these at finer spatial scales within cities is challenging, as, typically, surveillance of detected infections is aggregated at the city and country scales^5,13^.

Epidemiological models were informed with case count, mortality, and seroprevalence data to inform parameter estimation and describe the impact of NPIs on COVID-19 outcomes and community transmission of SARS-CoV-2^20^. Constrained epidemiological mathematical models allow the simulation of unobserved variables, such as the fraction of susceptible, susceptibility, and undetected infections. Mobile phone data measuring changes in transmission compared before the NPIs were included as co-variates, informing time-varying transmission rates that control some population-level effects of the NPIs. These estimates suggest that lockdowns successfully reduced community transmission, decreasing mortality and hospital admissions worldwide^21^. However, changes in mobility patterns depended on demographic and socioeconomic factors^2^, resulting in localized disease prevalence^1,22^.

Ideally, interventions would target the finest spatial scales: individuals and specific communities with a high frequency of contact. Indeed, individual-level interventions such as contact tracing were used to identify and break transmission chains, significantly reducing transmission^23^. However, only when a substantial portion of infections are ascertained and community transmission is low^24,25^. The high fraction of asymptomatic infections challenged the detection of SARS-CoV2 infections, the surveillance system’s testing capacity, and the time lags between exposure, symptom onset, and diagnosis (generation times)^25^. Consequently, the feasibility of contact tracing interventions is difficult in practice ^26–28^.

In addition, mobility data have been used in epidemiological models to inform meta-populations by simulating commuting between spatial units^29^. These studies concluded that commuter reductions matter, especially in the early epidemic stages, where infections are still sustained mainly by heterogeneous transmission events and not by average community infections ^30,31^. The role of mobility in altering transmission finer spatial scales is still not completely understood^18^. By assuming that the contact rate decayed proportionally to the mobility within a spatial unit, mobility data were used as covariates of transmission. This suggests that these changes are reasonable measures of social distancing in communities^32–34^. In big urban centers, statistical analyses have revealed the correlation between community transmission and a reduction in mobility^35,36^. Mobility indicators have also been used in statistical models to forecast COVID-19 incidence cases and mortality, outperforming their counterparts without mobility information, underscoring the importance of mobility delineating transmissions^35,37^. However, with explicit process-based models or methods to describe mechanisms, it is easier to quantify and document the effect of mobility transmission^25,29,34^. Specifically, accounting for the heterogeneity in human mobility, which is correlated with socioeconomic status (SES) and general health outcomes such as mortality due to SARS-CoV-2, is essential for localized interventions^1,5,7^.

The emerging spatial heterogeneity due to differential effects on mobility highlights the importance of understanding the effect of finer within-city NPIs that restrict human mobility in SARS-CoV-2 transmission. The effects of these NPIs could ease future mitigation and control of novel pathogen outbreaks, especially those with transmission modes like SARS-CoV-2. Evaluating and understanding the mechanisms behind less restrictive interventions, like the ones used in Bogotá, are relevant intervention options for future epidemics^38–42^.

In Bogotá, research teams in Colombia reported the state of the COVID-19 outbreak via epidemiological indicators such as the effective reproductive number (*Reff),* which quantifies community transmission and forecasts of incident-reported infections and mortality. Despite the availability of these epidemiological indicators, in Bogota, the NPIs were implemented in localities with a high rate of incident cases in the previous weeks. The rate of incident cases was defined as the number of cases divided by the census population in each locality. However, those estimates are typically reported at the administrative scales of cities, and their effect at finer spatial scales remains to be understood^43^. A better understanding would make these interventions feasible for curtailing an epidemic and causing fewer social and economic costs than other restrictions^39–41^. Our study combines population epidemiological analyses with mobile phone data to examine the consequences of localized interventions in Bogotá between July and August 2020. Our main objective is to quantify the effects of interventions on reducing community transmission and quantify changes in reported cases and mortality due to COVID-19. We additionally use movement data at the scale of the restrictions to generate hypotheses by analyzing mobility patterns and their role in determining the success of these interventions. Finally, we control for spatial heterogeneity in socioeconomic and demographic variables that impact those interventions.

## Materials and methods

### NPIs description

From July to September 2020, four rotational lockdowns targeting two regions of Bogotá were implemented. These geographically smaller lockdowns started on July 13th and ended on August 27th. In Figure 1, we present the localities in each group and the time of the restriction. In Figure 1C, we present maps highlighting the localities with curfews during each restriction in red.

### Data description

#### Mobility data

We used Facebook’s Data from the GeoInsights portal (https://dataforgood.facebook.com/dfg/tools), which consists of aggregate and anonymized Facebook app users’ locations. We used movement and population datasets to quantify the variation in mobility for each locality in Bogotá. For each locality, Facebook provides a baseline calculated as the average number of devices categorized as being in each location (population) or who had made a directional transition (commute) during the baseline period, conditional on the week and time of day.

#### Surveillance epidemiological data

We analyzed daily incident confirmed infections, aggregated based on the confirmation date (i.e., when individuals tested positive for SARS-CoV-2 as reported by relevant healthcare institutions), and mortality data due to COVID-19. The health secretary collected both surveillance streams at the individual level^44^. In the supplementary material (SM), Figure S1, we present the case data at the locality scale.

#### Socioeconomic data

We used demographic and socioeconomic information at the locality level, obtained from the National Administrative Department of Statistics (DANE) and socioeconomic data from the Colombia Multipurpose Survey (to obtain socioeconomic indicators to assess the confounding factors in the effectiveness of implemented lockdown policies). It combines elements of quality-of-life surveys to cover household expenditures, the labor market, and living conditions^45^. The data include census population data concerning the number of individuals, access to water and sewage systems, healthcare access, unemployment rates, access to piped water and electricity, and poverty levels. Table 2 includes a small description of the six variables and their units.

**Table 1.** Model comparison. Overall model comparison by WAIC

| Model | Explanatory variables used | WAIC |
| --- | --- | --- |
| 1 | Random | 1347152 |
| 2 | Random + Outside Movement | 1297152 |
| 3 | Random + Inside Movement + Outside Movement | 1246247 |
| 4 | Random + Inside Movement + Outside Movement + Number of people | 979988 |
| 5 | Random + Inside Movement + Outside Movement + Number of people + Aqueduct | 741807 |
| 6 | Random + Inside Movement + Outside Movement + Number of people + Aqueduct + Health care | 741805 |
| 7 | Random + Inside Movement + Outside Movement + Number of people + Aqueduct + Health care + Unemployment | 732633 |
| 8 | Random + Inside Movement + Outside Movement + Number of people + Aqueduct + Health care + Unemployment + Poverty | 747867 |

**Table 2.** Socioeconomic variables were used in the analyses.

| Variable | Description | Units |
| --- | --- | --- |
| Census population | Number of individuals in the locality. | Number of individuals |
| Healthcare access | Percentage of the census population who have access to healthcare. | Dimensionless |
| Access to water and sewage systems | Percentage of the census population who have access to sewage system. | Dimensionless |
| Unemployment rate | Number of individuals unemployed in the last year. | Number of individuals |
| Access to piped water and electricity | Percentage of the censed population that has access piped water and electricity. | Dimensionless |
| Poverty levels | Indicator of poverty calculated in the country. | Dimensionless |

### Description of methods

#### Quantifying community transmission

We quantified community transmission via surveillance of COVID-19 data for each locality in Bogotá (Supplementary Material Figure S1) by estimating the effective reproductive number (*R*_*eff*_). We used the R package **EpiNow2** ^46,47^, which uses the renewal equation for the *R*_*eff*_ estimation^47^. If conditions remain identical, the effective reproductive number quantifies the expected number of secondary cases arising from a person showing symptoms at a particular time. This indicator measures the instantaneous transmissibility of the virus. In addition, EpiNow2 estimates the delays from infection to symptom onset and testing to confirmation by diagnosis. The estimation thus controls for uncertainty in the report date and generation times^48^.

#### Mobility analysis

We investigated mobility patterns during the restrictions. We present changes in mobility during the lockdown periods by calculating the percentage change in mobility used as baseline mobility during the period before the interventions (one month before each restriction started). We distinguish between external (between localities) and internal (within localities) movements for each pair.

#### Counterfactual projections and inference

We used a deterministic epidemiological compartmental model informed by confirmed cases of COVID-19 and mortality data (Supplementary material section 2). We estimated the model parameters before the start of each lockdown (see Figure 1 for the period that each intervention spanned and Supplementary Material section 3). We simulated them forward in time to project the expected number of cases and deaths. Thus, we simulate a counterfactual scenario without a lockdown. We used the simulated data to estimate the effective reproduction number (*R*_*eff*_). We compared the changes in community transmission at the locality and city levels via the *R*_*eff*_. We also compared the changes in mortality and reported cases. We quantified the impact of these interventions on SARS-CoV-2 community transmission and COVID-19 mortality in Bogotá, Colombia, from March 20 to May 10, 2020. We subsequently extended our simulation to September 1, 2020. The compartmental model structure is presented in Supplementary Material Section 2 Figure S3. We model time-varying contact and detection rates, simulating contact changes during interventions and varying testing.

#### Statistical associations between transmission changes and sociodemographic factors

We investigate how mobility and socioeconomic factors (see the Mobility data and socioeconomic data section above) influence changes in the effective reproduction number (*R*_*eff*_). We used a spatiotemporal Bayesian hierarchical model^49^ to quantify the explanatory power of these data that mean weekly *R*_*eff*_. We investigated how *R*_*eff*_ it could be explained by sociodemographic factors spanning the study period between June 2020 and September 2020. Our Bayesian hierarchical model is a linear regression model that predicts the mean weekly *R*_*eff*_ for a total of T=18 weeks. Our linear model includes 9 possible spatiotemporal covariates, and we use information across the T=18 weeks and S=20 localities to estimate the model coefficients *X*_*jik*_ (j=1…,9 covariates, i=1…,18 weeks and j=1…,20 localities). The Supplementary material section 2 presents a mathematical description of the spatiotemporal Bayesian hierarchical model.

The hierarchical model implementation allows for the comparison of nested models, and we selected the best model via the stepwise backward variable selection approach using the Watanabe Akaike criterion (WAIC) to compare models and assess the predictive power of the variables included. Finally, we employed a simulation via the NIMBLE R package to establish 95% confidence intervals for the coefficient estimates ^50^. We calculated the confidence intervals for the fixed effects, assuming that the conditional modes of the random effects are fixed throughout the units.

## Results

We described changes in mobility patterns during the period of each restriction (Table 1 and Figure 1B) and distinguished between movement within localities (internal) and between localities (external). We found changes in internal and external localities depending on the timing restrictions. Figure 1B presents the changes in external and internal mobility over time, showing that changes in external mobility are more variable than internal mobility. We also find that the differences between the localities under lockdown and those without are more significant in the first rounds of lockdowns.

We differentiated four regions, each representing a distinct pattern in mobility dynamics. In Figure 1B, we present maps highlighting the intervened localities. The first restriction reduced commuters between localities (external mobility) but increased internal movements in the locked-down localities (internal mobility). In those without restrictions, two patterns appear: on some days, the external movement increased, and on others, it decreased; however, in both cases, the internal movement remained unchanged. The second round, the upper right plot, shows a similar pattern in the localities without restrictions: the external movement changed, and some days showed an increase, and some decreased, but the internal movement seemed minimally affected. However, restricted localities showed a decrease in external movement, although not as much as that observed in the first group.

We estimated the *R*_*eff*_ at the locality level using the reported COVID-19 infections to quantify community spread at this scale (see the section *titled “Quantifying community transmission* in Methods”). To investigate the effectiveness of this restriction, we present the distribution of the *R*_*eff*_ in each locality (see Figure 2) and highlight the periods corresponding to each lockdown (Figure 1B). In Figure 2, we also included community transmission in pre- and post-intervention. We find that the stringency of interventions varies across localities. First, some localities exhibit a significant reduction *R*_*eff*_ compared with the pre-intervention period. Localities including Antonio Nariño, Bosa, Rafael Uribe, Tunjuelito, Usme, and Kenedy quickly plateau in their *R*_*eff*_ reduction.

**Figure 2.**
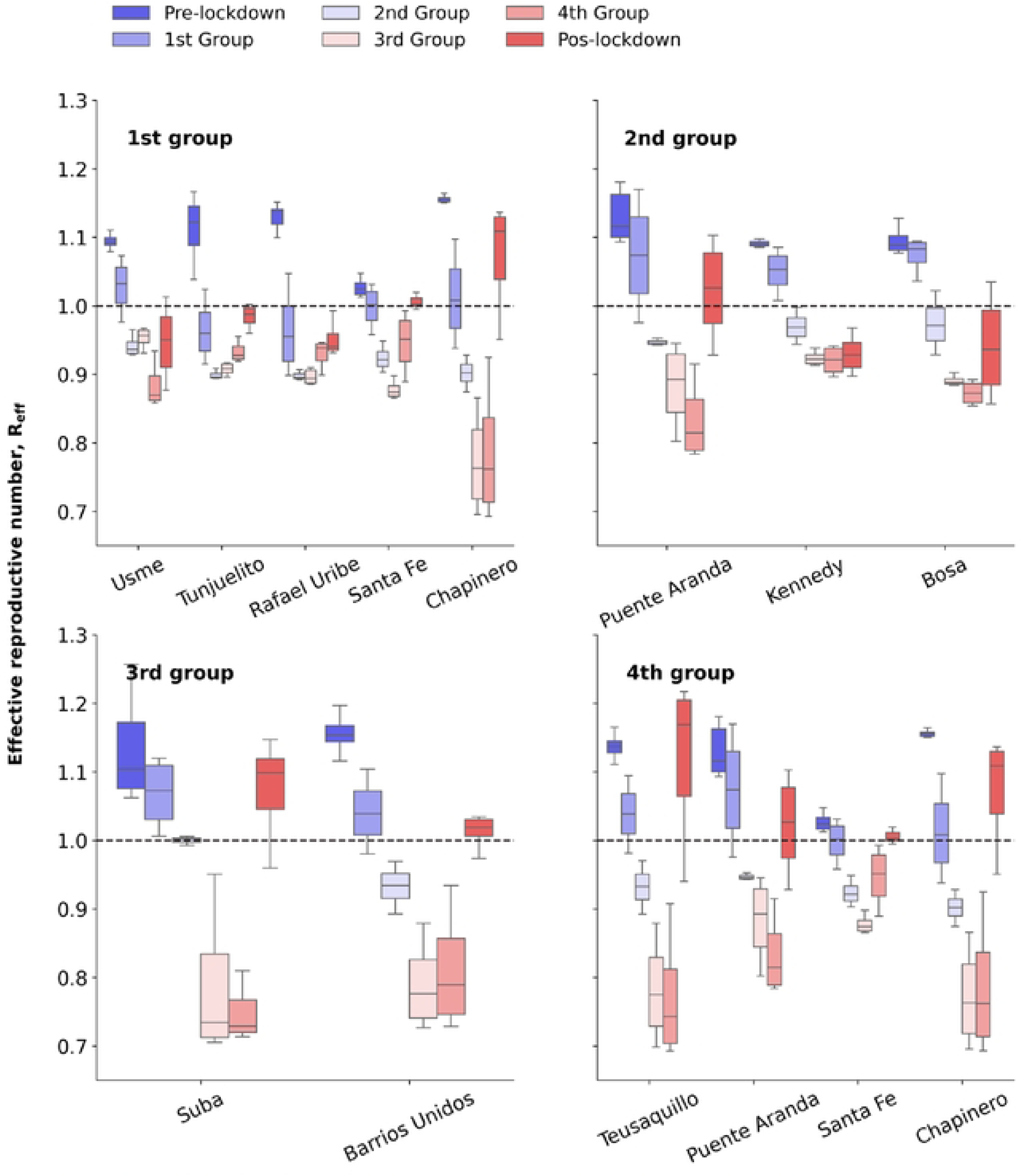
Spatiotemporal variation in the *R*_*eff*_ across localities. On the x-axis, we have every locality in the city; for each of those localities, we plot a box plot representing the median R_eff and the interquartile range for six periods before lockdown, four rotational lockdown rounds, and after lockdown for each locality in the city.

In contrast, other localities, including Chapinero, Suba Usaquen, and Teusaquillo, consistently decline *R*_*eff*_ as lockdowns are imposed (Figure 2). Together, these results show that finer lockdowns effectively reduced the community spread of SARS-CoV-2 at the city scale. However, these interventions are more effective in the first weeks of the restrictions (Figure 2). Because our community transmission estimates controls for susceptibility, this result points to a decline in compliance. As lower socioeconomic groups were restricted at first, the effect of the later interventions did not substantially affect transmission.

The counterfactual simulations of cases and mortality are presented in Figure 3. We found that the first round of lockdown reduced the number of detected incident cases of COVID-19 by 27% (95% Confidence interval CI, 22–36%). In the second, the model estimates the reduction was 19% (95% CI 17–22%). The third lockdown round impacted 11% (95% CI, 9–14%) of the detected infections. In addition, these rotational lockdowns led to reductions of 29%, 16%, and 8% in the number of deaths, respectively, for each round of intervention (as shown in Figure 3B). Figure 3C shows the simulation *R*_*eff*_ based on the observed data. Differences between the estimated and observed counterfactual are greater in the 3rd and 4th lockdowns.

**Figure 3.**
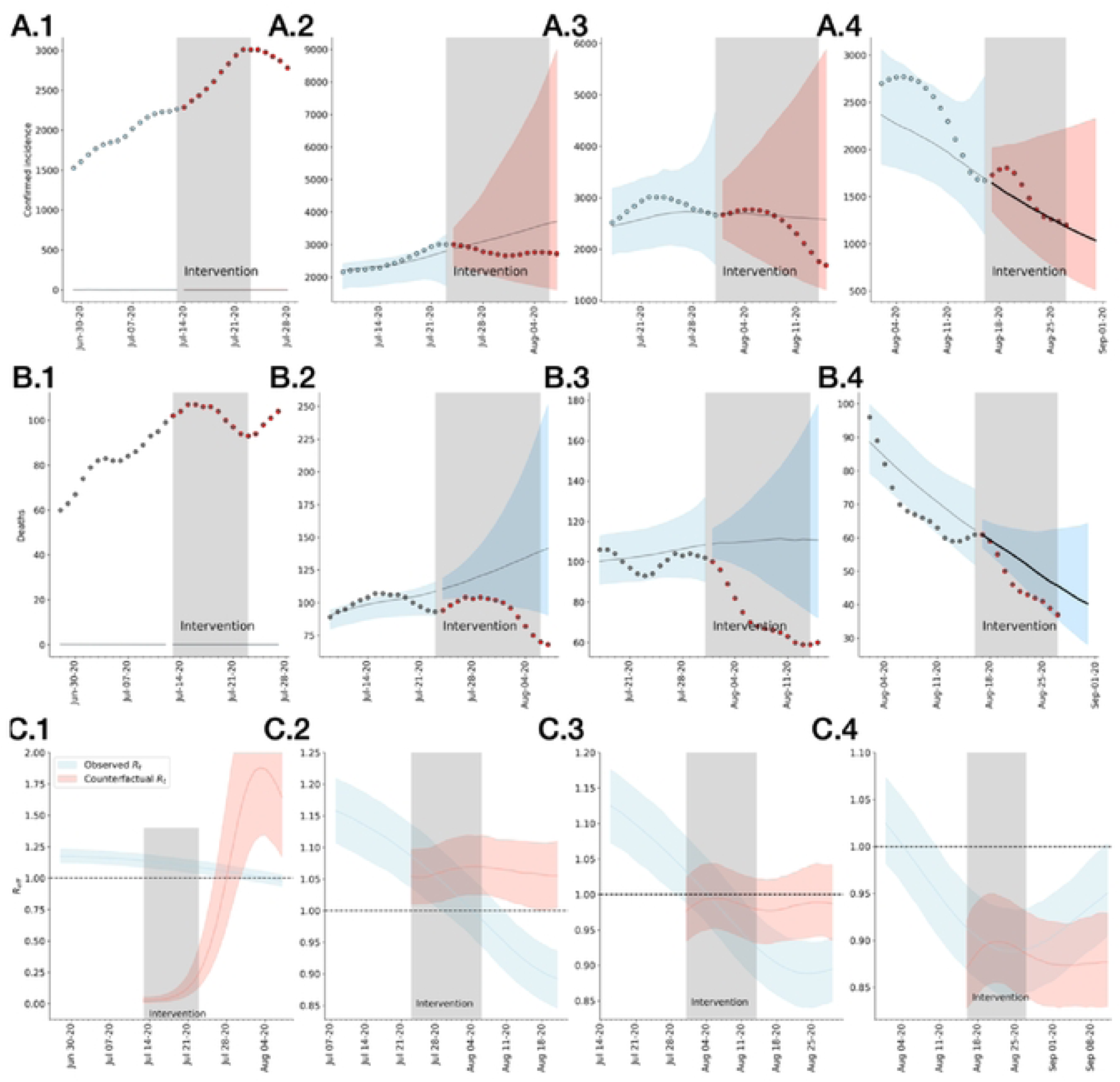
Counterfactual simulations. Control interventions starting two weeks earlier than the implemented measures commenced in early March. The simulations project Bogota’s daily cases and deaths via the model (depicted in panels A and B). In this figure, the solid line represents the median of the simulations, accompanied by their corresponding confidence intervals. The data points in the figures illustrate the observed cases and deaths during those periods. The upper row and lower row present counterfactual scenarios, with interventions being initiated on March 8 and March 1, respectively. The black lines, along with the surrounding bands, depict the median estimate, interquartile range, and 95% confidence intervals for these simulations.

To investigate how socioeconomic variables and population density are related to community transmission, we used a linear model to quantify whether changes in this data predict changes *R*_*eff*_ (see *the statistical model description* in the Methods section). Table 1 presents the Watanabe Akaike information criterion (WAIC) for the ten models. The best model includes mobility data, internal and external mobility, and socioeconomic data: population density, poverty, unemployment, and access to healthcare.

In Figure 4, we present mean parameter estimates of the best model, as well as uncertainty associated with the spatiotemporal variation (see Supplementary material figure S2). We find that localities that show a decrease in both internal and external mobility exhibit a lower effective reproduction number (*R*_*eff*_) (Supplementary material Table S1). We find that the localities that experienced greater reductions in *R*_*eff*_ had higher population density, higher electricity and piped water access, and lower unemployment rates (Figure 4). Similarly, the spatiotemporal model shows that reductions in *R*_*eff*_ correlated with localities with a low access rate to health care. Based on the model results, we identify that those reductions in mobility within and between areas explain changes in *R*_*eff*_ internal mobility, which, when reduced, contributes to a 16% decrease *R*_*eff*_ (95% CI, 12–19%). Regions with a larger average population size and density tend to exhibit smaller reductions in *R*_*eff*_. Changes in external mobility account for 17% of the variation (95% CI, 0% to 30%), and areas with lower unemployment rates account for 12% for a 95% confidence interval.

**Figure 4.**
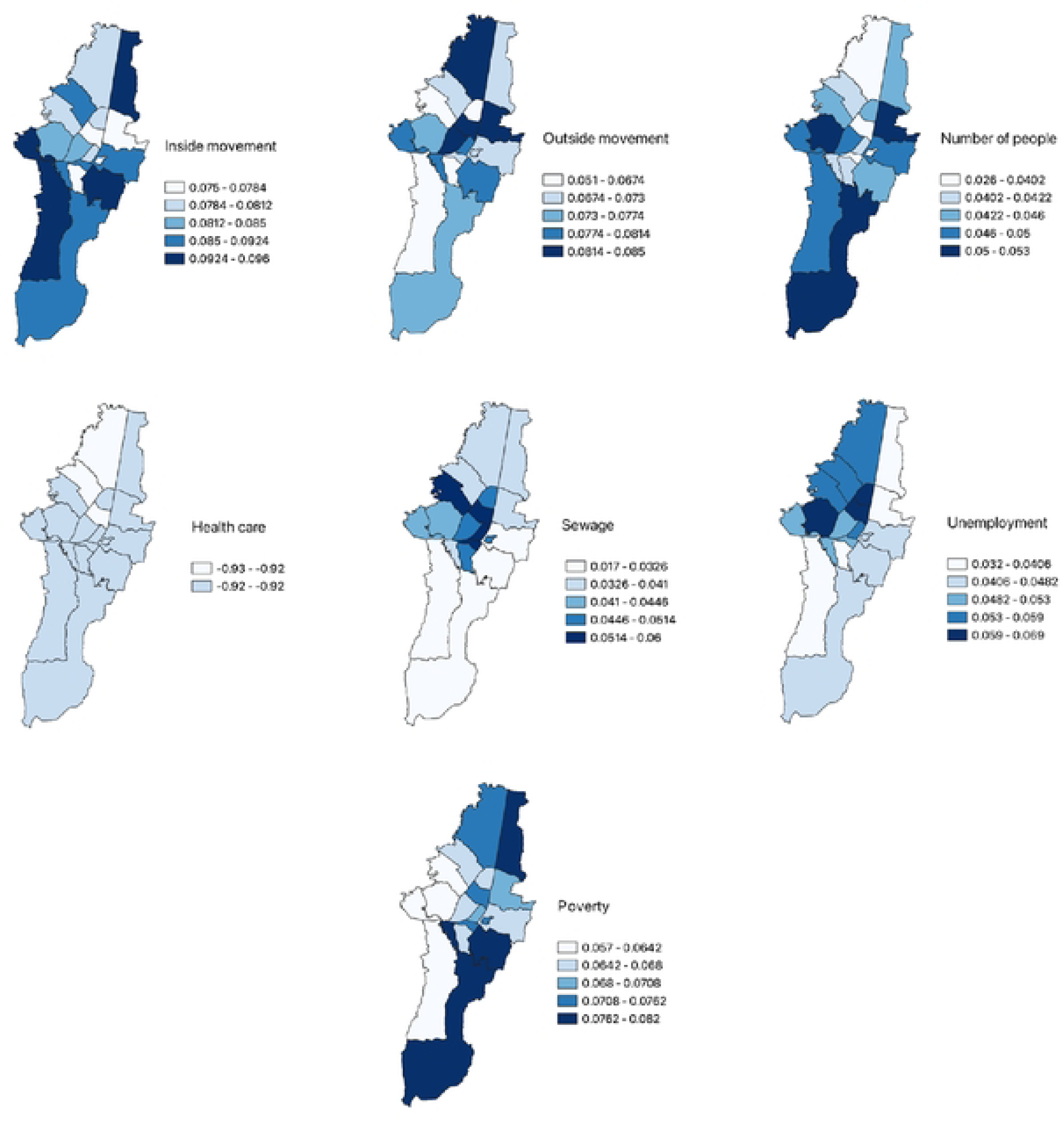
Parameter estimates for the best model. Estimated fixed effects (and 95% confidence intervals) for the model with the lowest Akaike information criterion (WAIC) heatmap for each locality of Bogotá. The fixed effects represent the estimated association between each independent variable and the Reff variation during the lockdowns, holding all else fixed.

## Discussion

In the absence of vaccines and other pharmaceutical interventions to treat and prevent infections and the transmission of communicable diseases, NPIs are the first control measure. These interventions, however, can tremendously impact communities relying on day-to-day economic activities, like the ones in LMICs. A reduction in mobility and, thus, contact by imposing curfews and city-level lockdowns successfully reduced SARS-CoV-2 activities upon introducing the virus to communities^26,35^. However, the success of finer interventions targeting specific communities or fields within city administrative aggregations remains to be further understood. It is challenging to quantify the effect of interventions at this level, mainly because cities are well-mixed places for individuals, especially in mega-cities. However, socioeconomic status has a clear spatial pattern, which is also related to population density and, thus, to the intensity of contacts that contribute to the transmission of these diseases. In this work, we describe a finer within-city lockdown implemented in Bogotá, D.C., the capital city of Colombia, an LMIC in Latin America. These NPIs were implemented to reduce hospital admissions due to COVID-19 during the first peak in Bogotá. Consequently, we do not need to control or discuss the presence of different virus strains or prior population immunity. Instead of reexamining the well-documented impact of mobility on disease transmission ^5,51^, this study focuses on how within-and between-unit mobility variations across localities affect the efficacy of lockdowns.

We used mobility data at the scale of the restrictions and showed that mobility was effectively reduced during the NPIs. However, the lockdowns were more effective in reducing the number of commuters between localities (external movement), while the within-localities movements were not diminished as equally (Figure 2). We used case count and mortality data of COVID-19 patients to inform epidemiological models and quantify community spread (*see Surveillance epidemiological data* section *in the Methods* section). We find that the effective reproductive number, or Reff, was diminished in all localities upon the start of the restrictions. Because each biweekly intervention was targeted at an aggregation of localities and the external mobility of the intervened spatial units was substantially diminished, our results suggest that a finer lockdown effectively reduced transmission via a reduction in mobility and, thus, contacts. However, because we do not know the mode of transportation of those commuters, it isn’t easy to understand the reason for the reduction in transmission. One transmission route could occur in public transit, as it is an important location where infected but asymptomatic individuals transmit the virus to those susceptible. Another is that commuters who work in supermarkets, grocery stores, and small shops are less frequently contacted by locked-down individuals. These can be infected and naive individuals who must acquire food and other supplies, contact infected commuters, and acquire the virus. In Bogotá, home delivery grocery shopping services exist, but their use is not popularized and likely depends on socioeconomic variables and age groups.

Motivated by the reduction in transmission across all localities and the reduction in mobility in a subset of the localities in the city, we simulated counterfactual scenarios to estimate changes in COVID-19 surveillance. The simulation assumed transmission did not change and was the same as the average of week before the lockdown started. We first showed that finer NPIs effectively reduced the *R*_*eff*_ and reduced the incident case counts and mortality data (Figure 2). Second, we found that the first intervention was the most effective in reducing transmission and resulted in the most significant reduction in cases and mortality. This result is interesting because it suggests that stringency to the NPIs across localities varied with time. Because the *R*_*eff*_ controls for the population’s susceptibility; those reductions in the effectiveness of the interventions across the study period are a consequence of increased contacts. Thus, we infer changes in compliance with the later interventions. However, from the mobility analysis, we did not observe a change in the mobility patterns during the later lockdowns (Figure 1). One possibility is that mobility data may be missing a fraction of the population, which is responsible for the inferred changes in compliance.

Previous theoretical simulations have suggested that lockdowns should be imposed for a substantial duration, such as 60 days, to contain the spread of a virus ^52^. Our results show that not only those localized interventions can slow community transmission but also that finer spatial scales impact the city-level transmission. However, we also show that controlling for spatial heterogeneity is necessary to account for factors such as poverty and population density, which contribute additional variation beyond what mobility explains.

We also find that these reductions were greater in localities with high socioeconomic status (SES). A reduction in mobility was also associated with localities with higher SES. Together, these results suggest that mobility, both internal and external movement between localities, is an important driver of community transmission. In contrast, internal mobility varies considerably across lockdown rounds, which can be explained by differences in demographics and socioeconomic status (SES). For example, reducing activities away from home while increasing the time spent at home depends on economic conditions and time-use patterns. This pattern is evident in our results, as reductions in transmission in each locality are heterogeneous. This heterogeneity can be explained by the fact that these restrictions are linked to population density and SES, which modulate their potential to slow new infections by restricting geographical mobility. For example, restrictions in localities that result in workplace closures and limits on gatherings could generate more significant reductions in *R*_*eff*_ than those in residential units.

The impact of COVID-19 in Bogotá varies across different neighborhoods, with areas of lower socioeconomic status facing a more significant burden. Research shows that these communities struggle to adhere to public health measures, leading to an increase in the number of COVID-19 cases. A study demonstrated a rebound effect on the adequate reproductive number after quarantine shifts, particularly in areas such as Candelaria, Chapinero, Suba, Teusaquillo, and Usaquén, possibly due to economic density. Research has emphasized the connection between socioeconomic status and mobility response to the pandemic, indicating that higher SES is associated with a more significant decrease in mobility^1,7^. Our findings suggest that changes in disease transmission are less pronounced in households from neighborhoods with lower socioeconomic status. In cities such as Bogota, where significant inequalities exist, the impact of the COVID-19 pandemic has amplified the challenges faced in socially divided urban areas. These disparities highlight the importance of recognizing different responses to policy measures to guide targeted public investments and government interventions, particularly during critical periods that may require new or localized lockdowns.

This approach allows us to assess how variations in transmission intensity, influenced by mobility, demographic characteristics, and socioeconomic status (SES), affect Reff. Our methodology integrates mobility data with socioeconomic and demographic datasets to quantitatively analyze the relationships between effective reproduction number change (Reff) changes and these covariates. The results from our linear mixed-effects model highlight the importance of demographic, economic, and mobility-related conditions in shaping Reff during interventions in Bogotá.

Our work suggests that controlling for socioeconomic heterogeneities is of interest when designing NPIs. Importantly, we show that, within city, NPIs effectively reduce city-scale community transmission. Thus, exploring finer and customized interventions will help protect people’s health and reduce future economic kicks. The approach outlined in the study prioritizes the reopening of the intervened spatial scale based on their risk profiles. The risk, however, was defined on cumulative data and not any intelligent community transmission index. Investigating if refining this indicator could have revealed a different intervention scheme would be interesting. Specifically, implementing a small-area lockdown strategy in Bogotá requires strong surveillance capabilities, early warning systems, and rapid response mechanisms to manage surges in case transmission effectively. Without these essential components, the strategy may be ineffective. While the evaluation focuses on short-term outcomes, it remains uncertain whether this approach to social distancing can be sustained and proven effective in controlling the epidemic curve in the long run. Learning from experiences in other countries, we learned that prematurely lifting school closures could increase urban mobility and potentially impact disease transmission. Therefore, any decisions regarding school closures must be carefully planned to mitigate the risk of a resurgence of the outbreak.

While we controlled for spatial heterogeneities encoding socioeconomic variability that are suspected of stratifying mobility changes during the restrictions, we are not concluding causality. We limited our methodology to summarize and regress socioeconomic data and mobility to explain changes in transmission. A significant source of variability that remained uncontrolled is the compliance to interventions as a function of the awareness of disease and transmission. Thus, changes in human behavior, even in the absence of government interventions, should impact community transmission. As a result, it is likely that during the peak, protecting behaviors emerged and reduced transmission, and thus, suggest we overestimate simulated counterfactual data. Another potential limitation is we assume stationary infection fatality rates, which were known to increase during the COVID-19 peak in Colombia and some countries in the US. Increases result in higher fatalities, suggesting we could underestimate these in our projections.

Future research could investigate how spreading within specific communities influences behavior not captured by movement data. Additionally, while our estimates control for varying population susceptibility at the level of the restrictions, this is likely clustered in finer spatial scales related to socioeconomic variables. Thus, compliance at a finer spatial scale could be a factor that explains our inference that later interventions were not as effective as the first ones.

## Data availability

The data for this paper are available at this link.

## Code availability

The code is available in this GitHub repository.

## Data Availability

Data will be available in a public repository

## Acknowledgments

We thank the Instituto Nacional de Salud for providing the COVID-19 incidence data. The study was funded by a mixed-methods study on the design of AI and data science-based strategies to inform public health responses to COVID-19 in different local health ecosystems within Colombia (COLEV) project funded by the International Development Research Centre (IDRC) and the Swedish International Development Cooperation Agency (SIDA) [109582]. We thank Alejandro Feged and Felipe Gonzalez for providing the mobility data for the analysis.

## Ethics declarations

